# Ethnicity, comorbidity, socioeconomic status, and their associations with COVID-19 infection in England: a cohort analysis of UK Biobank data

**DOI:** 10.1101/2020.05.06.20092676

**Authors:** Albert Prats-Uribe, Roger Paredes, Daniel Prieto-Alhambra

## Abstract

**Objectives:** Recent data suggest higher COVID-19 rates and severity in Black, Asian, and minority ethnic (BAME) communities. The mechanisms underlying such associations remain unclear. We aimed to study the association between ethnicity and risk of COVID-19 infection and disentangle any correlation with socioeconomic deprivation or previous comorbidity.

**Design:** Prospective cohort.

**Setting:** UK Biobank linked to Hospital Episode Statistics (HES) and COVID-19 tests until 14 April 2020.

**Participants:** UK Biobank participants from England, excluding drop-outs and deaths.

**Main measures:** COVID-19 infection based on a positive PCR test. Ethnicity was self-reported and classified using Office of National Statistics groups. Socioeconomic status was based on index of multiple deprivation quintiles. Comorbidities were self-reported and completed from HES.

**Analyses:** Multivariable Poisson analysis to estimate incidence rate ratios of COVID-19 infection according to ethnicity, adjusted for socioeconomic status, alcohol drinking, smoking, body mass index, age, sex, and comorbidity.

**Results:** 415,582 participants were included, with 1,416 tested and 651 positive for COVID-19. The incidence of COVID-19 was 0.61% (95% CI: 0.46%-0.82%) in Black/Black British participants, 0.32% (0.19%-0.56%) in ‘other’ ethnicities, 0.32% (0.23%-0.47%) in Asian/Asian British, 0.30% (0.11%-0.80%) in Chinese, 0.16% (0.06%-0.41%) in mixed, and 0.14% (0.13%-0.15%) in White. Compared with White participants, Black/Black British participants had an adjusted relative risk (RR) of 3.30 (2.39-4.55), Chinese participants 3.00 (1.11-8.06), Asian/Asian British participants 2.04 (1.36-3.07), ‘other’ ethnicities 1.93 (1.08-3.45), and mixed ethnicities 1.07 (0.40-2.86). Socioeconomic status (adjusted RR 1.93 (1.51-2.46) for the most deprived), obesity (adjusted RR 1.04 (1.02-1.05) per kg/m2) and comorbid hypertension, chronic obstructive pulmonary disease, asthma, and specific renal diseases were also associated with increased risk of COVID-19.

**Conclusions:** COVID-19 rates in the UK are higher in BAME communities, those living in deprived areas, obese patients, and patients with previous comorbidity. Public health strategies are needed to reduce COVID-19 infections among the most susceptible groups.

## Background

By the end of 22 April 2020, illness caused by SARS-CoV-2 infection, also known as COVID-19, had been diagnosed in 133,495 people and killed 18,100 people in the United Kingdom (UK). As the virus spreads nationally and internationally, data continue to arise on subpopulations most at risk of contracting this disease and developing its severest forms. Weekly data from the UK Office for National Statistics (ONS) [1] suggest that COVID-19 was responsible for 6,213 deaths in England and Wales in the week ending 10 April 2020, equivalent to 33.6% of all deaths observed. Although almost 40% of the casualties were patients aged 75 to 84 years old, there is increasing evidence of life-threatening forms of COVID-19 in younger age groups. Almost 5% of COVID-19-related ICU admissions in Italy have been under 40 years old, and another 36% have been 40 to 60 years old. Overall, 82% of these patients have been men. [2]

Concerns have been raised recently that Black, Asian, and minority ethnic (BAME) communities in the UK might be at higher risk of COVID-19 mortality, including BAME healthcare professionals. [3] There is, however, a scarcity of data on whether this excess mortality is related to higher susceptibility to infection, higher risk of lethal forms of the disease, or poorer prognosis. Emerging data from seroprevalence studies [4] and US hospital electronic medical records [5] do not support these hypotheses, suggesting geographic heterogeneity.

Preliminary data also suggest a higher rate of infections in low-income households [6] and greater severity and rates of complications and death in older people, men, and people with specific comorbidities, such as hypertension.[7–9] Understanding the association between socioeconomic deprivation and the risk of COVID-19 infection is key to devising targeted, efficient interventions to limit its transmission in the community.

We obtained data from the UK Biobank, a large UK cohort, to unravel the associations between ethnicity, socioeconomic status, and the risk of COVID-19 infection.

## Methods

### Study design and setting

We used a prospective cohort study. All participants who were registered in UK Biobank [10] and living in February 2018 and who had not withdrawn permission to use their data by 7 February 2020 were included. Those residing in Scotland, Wales, or Northern Ireland were excluded as test data were only available for England.

The UK Biobank study recruited more than 500,000 participants from 37 to 73 years old through postal invitation to those who are registered with the UK National Health Service, living in England, Scotland, and Wales. Demographics, lifestyle, disease history, and physiological measurements were collected via questionnaires, physical measurements and interviews in baseline assessments (2006-10). [10,11] Participants gave informed consent for data linkage to medical records. The participants tend to be healthier than the general UK population. [12]

Participants were followed-up using COVID-19 test data from 16 March 2020 to 14 April 2020. Data was linked to COVID-19 tests (see Study Outcome) and hospital inpatient data from Hospital Episode Statistics (HES).

### Study outcome

The main outcome was SARS-CoV-2 infection based on a PCR test. This information was retrieved from UK Biobank linkage to Public Health England COVID-19 test data.[13] We obtained data on who was tested, test results, and whether the test sample was taken in an inpatient setting. Patients were considered positive if one or more of the tests performed were positive for SARS-CoV-2. Participants were considered inpatients if one of their samples was marked as from an inpatient setting.

### Exposures and measurements

The main exposure variable was self-reported ethnicity, collected during the participant’s UK Biobank recruitment visit using a touch-screen questionnaire. This information was classified using Office of National Statistics groups into Asian/Asian British, Black/Black British, Chinese, mixed, White, and other groups. “Prefer not to answer” and “Don’t know” responses were grouped together.

Socioeconomic status was based on the index of multiple deprivation (IMD) collected at recruitment into the UK Biobank. IMD England 2010 index, rank, and deciles were used to stratify participants into IMD quintiles.

Sex and year of birth were acquired from the National Health Service Central Register (NHSCR) at recruitment, but in some cases were updated by the participant. BMI was calculated by the UK Biobank using physical measurements collected at the recruitment visit. Alcohol drinking and smoking were self-reported during the same appointment.

Where information on ethnicity, IMD, sex, year of birth, BMI, alcohol use, or smoking were missing at baseline, information collected at the further 3 visits was used to maximise completeness.

We selected the comorbidities that are most likely to be risk factors for COVID-19 based on previous literature [14,15] and assessed them using pre-specified ICD-10 codes from linked HES data from the end of March 1992 to the end of March 2017. We included hypertensive disease (I10-I15), diabetes mellitus (E10-E14), ischaemic heart diseases (I20-I25), other forms of heart disease including hearth failure (I30-I52), chronic lower respiratory diseases including COPD and asthma (J40-J47), and renal failure (N17-N19).

This research was conducted using the UK Biobank Resource under Application Number 46228. Although the original application was unrelated to COVID-19 work, an exception was made to allow these linked data to be used for COVID-19 research without further applications, to maximise the speed of the proposed study [16].

### Statistical analysis

We calculated the proportion or mean and standard deviation of each exposure variable for the population as a whole and stratified by test status (not tested, tested, and tested positive). We computed cumulative incidence of testing and positive tests for each ethnic background group and IMD quintile. We fitted a multivariable Poisson model to estimate the risk ratios of SARS-CoV-2 infection according to ethnicity, socioeconomic status, and the presence or absence of the included comorbidities. All models were further adjusted for age, sex, alcohol drinking, smoking, and body mass index (BMI). Patients with missing data in any regressor were excluded from the multivariable regression. We did not test for interactions or perform subgroup analyses due to limited statistical power.

Data management was performed in Python 3.7.6.[17] All analyses were performed in STATA version 15.1.[18]

## Results

After excluding 69,773 people due to country of residence, 17,151 due to death, and 13 due to opt outs, we included 415,582 UK Biobank participants in our sample. Of these, 1,416 (0.34%) had been tested and 651 (0.16%) had tested positive for COVID-19 infection by 14 April 2020. Detailed baseline characteristics of all participants and stratified by COVID-19 testing and infection status are reported in Table 1.

**Table 1.** Sociodemographic information and self-reported history of illness of UK Biobank participants, stratified by COVID-19 testing status. Values are percentages for categorical variables and mean (SD) for body mass index (BMI).

|  | Total | Not tested | Tested | Tested positive |
| --- | --- | --- | --- | --- |
|  | *N* = 415,582 | *N* = 414,166 | *N* = 1,416 | *N* = 651 |
| **Age categories** |  |  |  |  |
| 49-54 | 6.00% | 6.10% | 5.90% | 5.80% |
| 55-59 | 12.80% | 12.80% | 13.20% | 14.90% |
| 60-64 | 14.60% | 14.60% | 11.30% | 10.80% |
| 65-69 | 16.90% | 16.90% | 12.60% | 13.10% |
| 70-74 | 23.50% | 23.50% | 20.10% | 19.50% |
| 75-79 | 20.10% | 20.00% | 26.30% | 25.00% |
| 80-86 | 6.10% | 6.10% | 10.70% | 10.90% |
| Missing | 0% | 0% | 0% | 0% |
| **Sex** |  |  |  |  |
| Female | 55.00% | 55.00% | 46.70% | 43.60% |
| Male | 45.00% | 45.00% | 53.30% | 56.40% |
| Missing | 0% | 0% | 0% | 0% |
| **Alcohol use** |  |  |  |  |
| Never | 4.40% | 4.40% | 5.80% | 6.10% |
| Previous | 3.40% | 3.40% | 5.60% | 6.00% |
| Current | 91.80% | 91.80% | 88.10% | 87.30% |
| Prefer not to answer | 0.10% | 0.10% | 0.20% | 0.10% |
| Missing | 0.20% | 0.20% | 0.20% | 0.50% |
| **Smoking** |  |  |  |  |
| Never | 55.20% | 55.10% | 45.10% | 45.80% |
| Previous | 34.10% | 34.40% | 40.00% | 42.20% |
| Current | 9.90% | 9.90% | 14.10% | 10.60% |
| Prefer not to answer | 0.40% | 0.40% | 0.60% | 0.90% |
| Missing | 0.20% | 0.20% | 0.20% | 0.50% |
| **BMI (mean (SD))** | 27.4 (4.8) | 27.4 (4.8) | 28.7 (5.7) | 29.2 (5.5) |
| Missing | 0.57% | 0.56% | 1.70% | 1.49% |
| **Index of multiple deprivation (IMD) quintiles** |  |  |  |  |
| 1 (most affluent) | 29.60% | 29.60% | 22.30% | 20.00% |
| 2 | 23.80% | 23.90% | 17.70% | 15.50% |
| 3 | 17.90% | 17.90% | 18.60% | 19.80% |
| 4 | 15.70% | 15.70% | 19.10% | 20.40% |
| 5 (most deprived) | 13.00% | 13.00% | 22.40% | 24.30% |
| Missing | 0% | 0% | 0% | 0% |
| **Comorbidities** |  |  |  |  |
| I10-I15 Hypertensive diseases | 22.47% | 22.41% | 39.12% | 38.56% |
| E10-E14 Diabetes mellitus | 5.92% | 5.89% | 14.05% | 16.13% |
| I20-I25 Ischaemic heart diseases | 8.11% | 8.08% | 16.95% | 15.67% |
| I30-I52 Other forms of heart disease | 7.78% | 7.74% | 18.79% | 17.05% |
| J40-J47 Chronic lower respiratory diseases | 9.46% | 9.43% | 18.08% | 17.51% |
| N17-N19 Renal failure | 2.58% | 2.55% | 10.31% | 9.68% |

Tested and COVID-19 infected participants were older and had a higher proportion of men than non-tested participants. Mean (standard deviation (SD)) BMI was higher among tested (28.7 (5.7) kg/m2) and COVID-19 infected (29.2 (5.5)) participants than non-tested participants (27.4 (4.8)). Almost twice the proportion of tested (21.5%) and infected (23.6%) participants lived in the most deprived areas than non-tested participants (12.3%). Similarly, participants infected with COVID-19 had a higher prevalence of comorbidity than non-tested participants, including hypertension (38.6% infected vs 22.4% non-tested), diabetes (16.1% vs 5.9%), ischaemic heart disease (15.7% vs 8.1%), other cardiopathies (17.5% vs 7.7%), chronic respiratory disease (17.5% vs 9.4%), and renal failure (9.9% vs 2.5%).

Table 2 reports the incidence of testing and testing positive for COVID-19 overall and for inpatient tests, stratified by ethnic background. Ethnicity was missing for 764 (0.20%) participants. There were clear differences in testing rates, with a higher incidence of tests among Black/Black British (0.89% (95% CI: 0.70%-1.14%)), other ethnic group (0.62% (0.42%-0.92%)), and Asian/Asian British (0.53% (040.%-0.70%)) participants than among Chinese (0.30% (0.11%-0.80%)), mixed (0.27% (0.13%-0.57%)), and White (0.32% (0.31%-0.34%)) participants. Similar patterns were seen when only inpatient testing was considered.

**Table 2.**
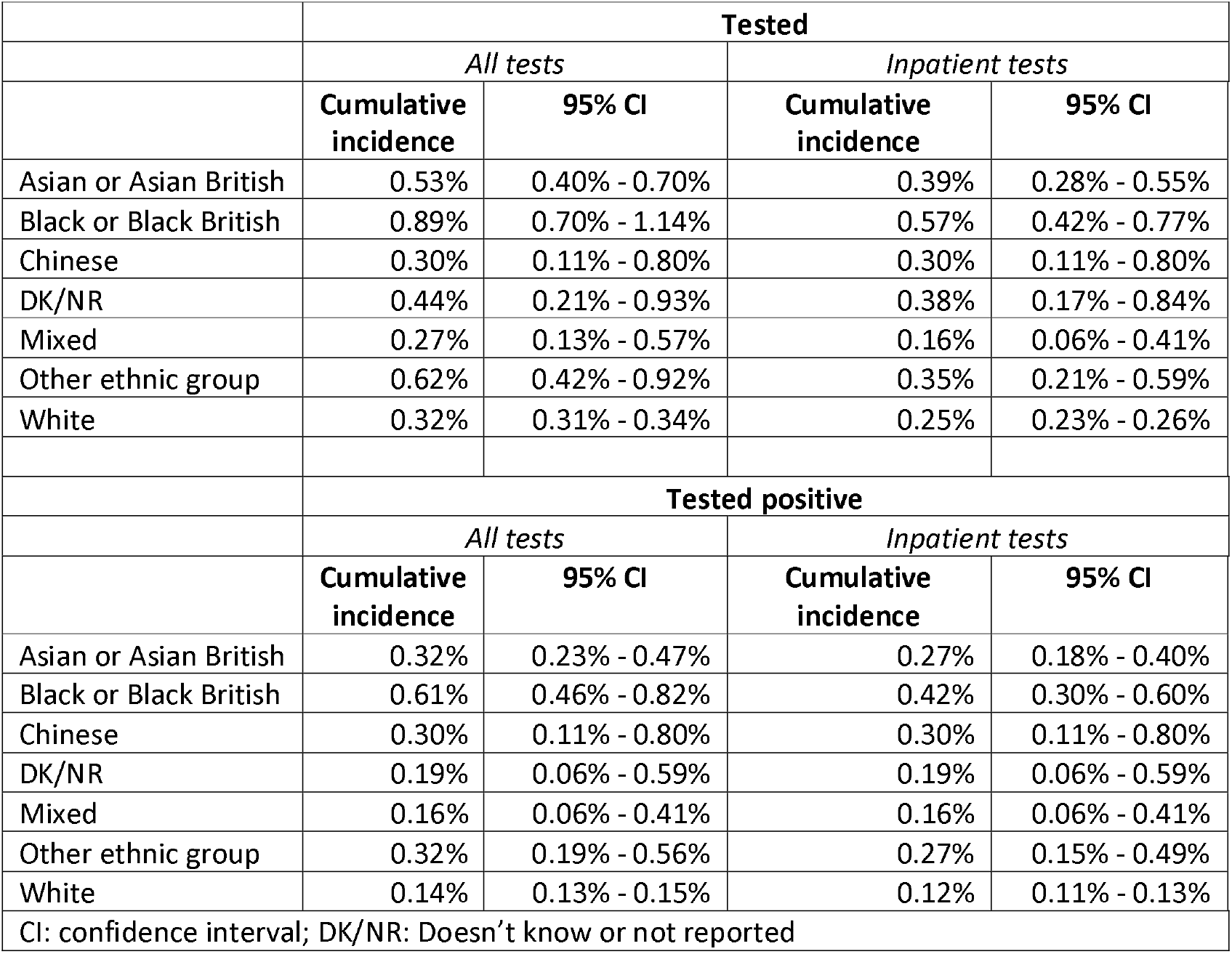
Cumulative incidence (and 95% confidence interval) of those tested and tested positive for COVID-19 infection between 16 March 2020 and 14 April 2020, stratified by ethnic background and where the test was performed.

There were also noticeable disparities in the incidence of COVID-19 infections according to ethnicity, again with the highest rates among Black/Black British participants (0.61% (0.46%-0.82%)) and the lowest rates among White participants (0.14% (0.13%-0.15%)). Again, similar patterns were observed when the analysis was restricted to inpatient diagnoses.

Participants with any missing information were excluded from further analysis, excluding 2,815 (0.68%).

The observed differences were not attenuated after multivariable adjustment for age, sex, BMI, smoking, alcohol drinking, socioeconomic status, and comorbidity (Figure 1). Compared to White participants, Black/Black British participants had an adjusted relative risk (95% CI) for COVID-19 infection of 3.30 (2.39-4.55), other ethnic groups a risk of 1.93 (1.08-3.45), Asian/Asian British participants a risk of 2.04 (1.36-3.07), and Chinese participants a risk of 3.00 (1.11-8.06). Participants with mixed ethnicity (1.07 (0.40-2.86)) did not differ from White participants.

**Figure 1.**
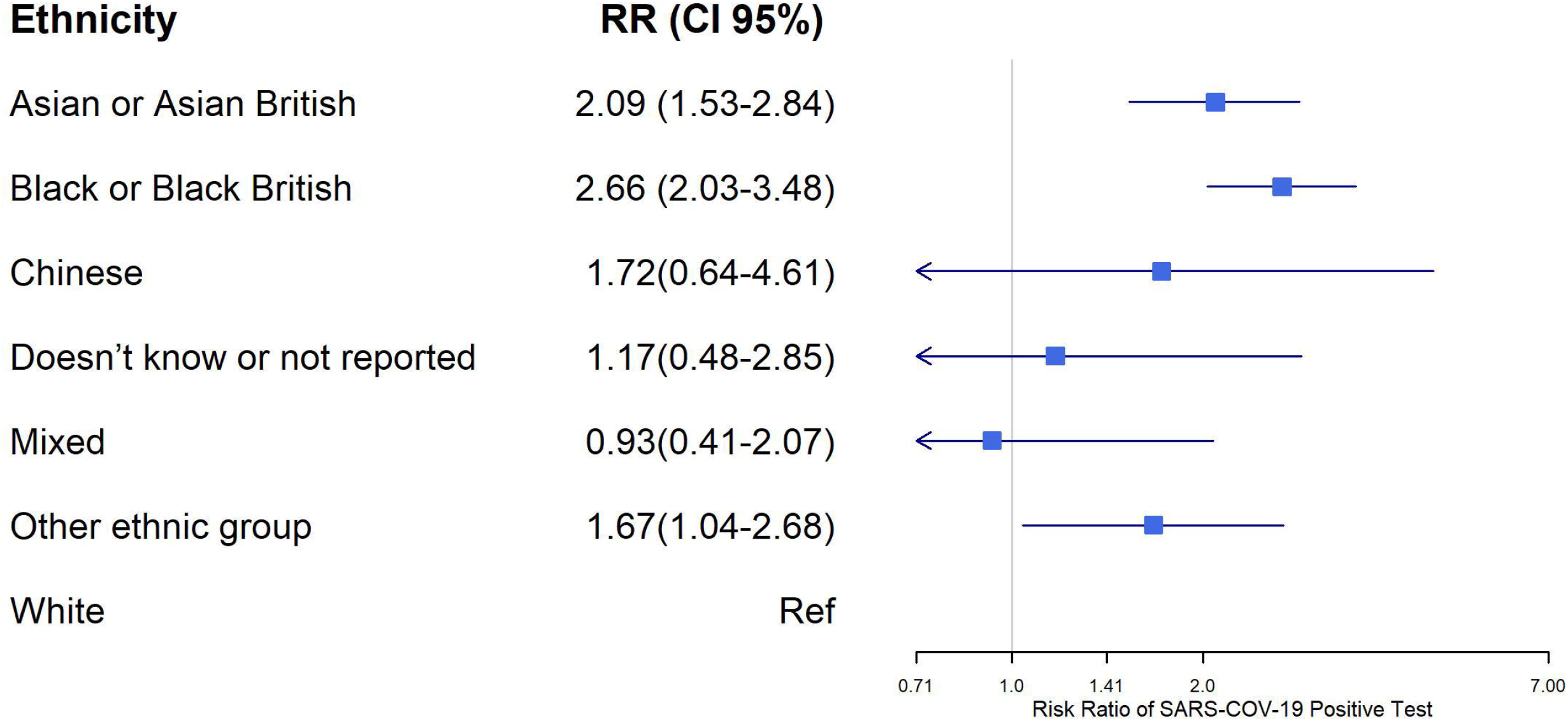
Adjusted relative risks of COVID-19 infection according to ethnicity

Multivariable analysis showed that socioeconomic deprivation and comorbidity were independently associated with an increased risk of COVID-19 infection (Figure 2). Using the first IMD quintile (least deprived) as the reference group, the adjusted relative risk of infection increased monotonically with IMD quintile, reaching almost double the risk in the fifth, most deprived quintile (relative risk: 1.93 (95% CI: 1.08-3.45). All of the tested comorbidities except history of ischaemic heart disease were independently associated with an increased risk of COVID-19 infection (history of hypertension, non-ischaemic heart disease, chronic respiratory disease, and renal impairment).

**Figure 2.**
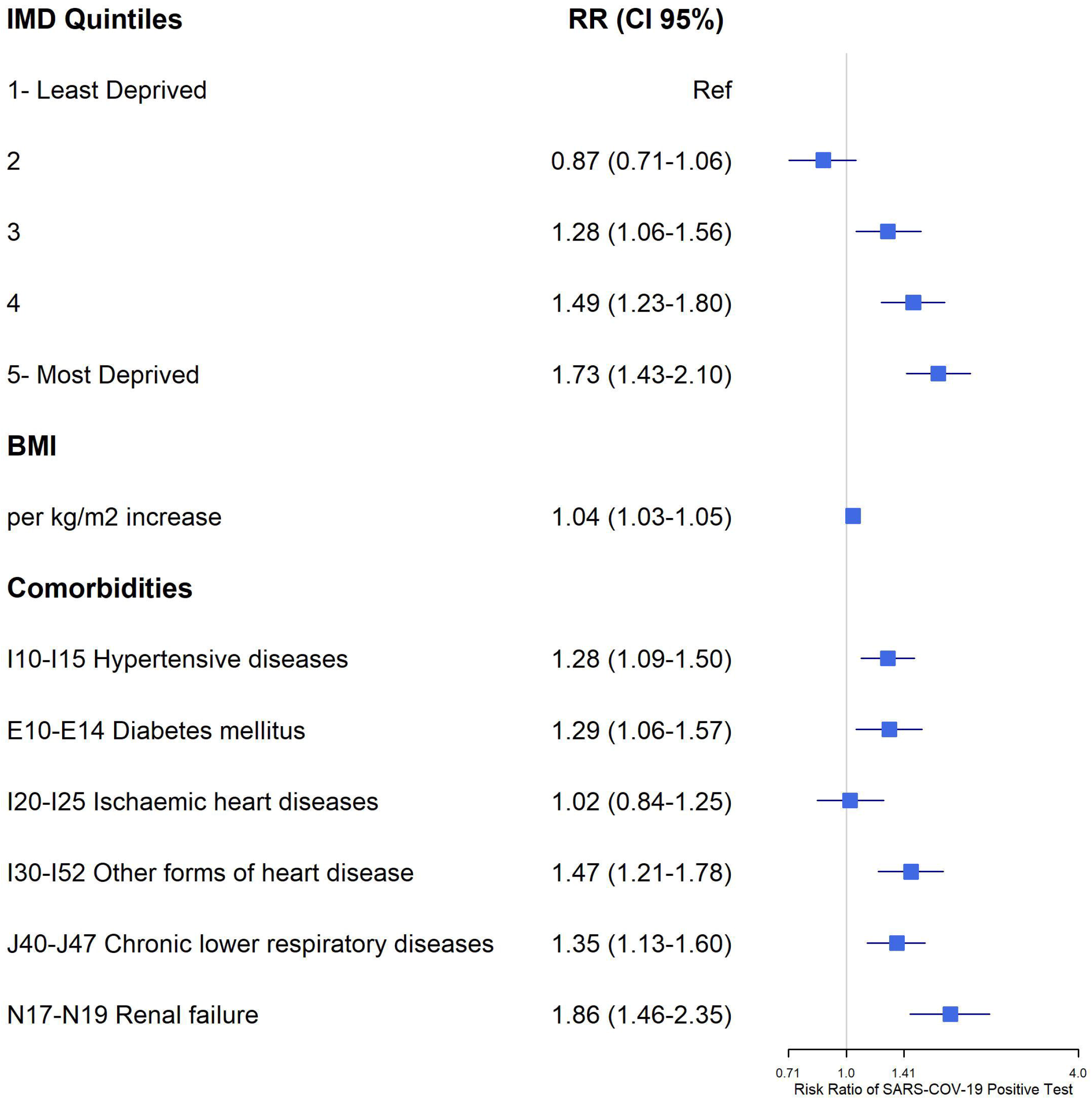
Adjusted relative risks of COVID-19 infection according to socioeconomic status (Index of Multiple Deprivation (IMD) quintiles) and comorbidity

High body weight was also independently associated with COVID-19 infection risk, with an adjusted relative risk of 1.04 (1.02-1.05) per kg/m^2^ increase in BMI. We did not find an association with self-reported alcohol drinking or smoking status (data not shown).

## Discussion

To our knowledge, this is the first study of the association between sociodemographics, ethnicity, and COVID-19 infection risk in the UK. In a cohort of over 415,000 participants, BAME communities appeared to be at higher risk of COVID-19 infection. Asian/Asian British and ‘other’ ethnic group participants had approximately double the risk of White participants, while Chinese and Black/Black British participants had more than triple the risk. This association was independent of sex, age, BMI, lifestyle risk factors, socioeconomic status, and comorbidity.

Despite a number of initial reports declaring differences in genetic predisposition to COVID-19 according to ethnicity, this difference has been disproved more recently as the virus has spread globally and people of all ethnic groups have been affected. [19] Data from previous pandemics, like the 2009-2010 H1N1 influenza, suggest that differences in preventive behaviours might explain the discrepancy in the spread of airborne viral infections across different ethnic groups.[20] As uncertainty remains as to the mechanisms behind the observed increase in risk of COVID-19 infection and mortality among BAME groups, recent reports have called for a National Commission on COVID-19 Racial and Ethnic Health Disparities in the US. [21] Similar requests have been made by a number of bodies to the UK government.[3]

The observed association between socioeconomic deprivation and the risk of COVID-19 infection is also novel. A clear “dose-response” effect was seen, with those living in the third, fourth, and fifth most deprived areas of the country having a 58%, 63%, and approximately 100% greater risk of contracting COVID-19, compared with those in the least deprived areas of the country. Similar associations have been reported for other pandemics in the UK.[22]

We can only speculate on the mechanisms underlying the observed association between socioeconomic status and COVID-19 risk. Income and employment deprivation, which are major contributors to the IMD, may limit people’s ability to make informed, free choices about self-protection. People with low or less stable income might be forced to choose jobs with higher intrinsic risk of contagion. Barriers to housing and services can lead to overcrowded households and environments, where transmission is more likely to occur and suggested self-isolation procedures are impossible to follow. People in more deprived areas may have less access to healthcare, and worse healthcare resource availability has been shown to lead to an increase in COVID-19 mortality. [23] Research on the social determinants of COVID-19 is urgently needed.

Our study confirms previous findings on common comorbidities associated with COVID-19, including hypertension, diabetes, overweight/obesity, heart disease, chronic respiratory disease, and renal impairment. In a recent study of 5,700 participants, [14] the most common comorbidities observed among people admitted with COVID-19 to 12 hospitals in New York City were hypertension (56.6%), obesity (41.7%), and diabetes (33.8%). Similar findings have been reported elsewhere. [15]

Our study has several limitations. We did not have data on region of residence. According to ONS, [1] there are geographic differences in mortality attributable to COVID-19 in the UK, with the highest rates currently in London and the West Midlands. Heterogeneous distribution of ethnic groups and socioeconomic status across the country could partially explain the observed effect.

Misclassification of COVID-19 infection due to differential testing across ethnic or socioeconomic strata may have affected the accuracy of our results. However, our data were collected when only severe cases were tested, and our findings were similar when restricted to inpatient testing, suggesting that even with these potential misclassifications, the observed associations remain clinically relevant. There was also potential for misclassification of comorbidity. However, we used linked electronic medical records to minimise recall bias and collect the most recent comorbidities.

The observational nature of this study makes the results susceptible to confounding and prevents us from inferring causality. We also lacked power to explore interactions or perform stratified analyses between the studied socioeconomic factors. There is a need to further explore the synergistic effects between these and with other health determinants on COVID-19 risk.

This study also has strengths, particularly surrounding the data used. We used a large sample with little missing data, links to hospital data, and self-reported information, which is considered a reliable source for ethnicity.[24] We also had very reliable information on COVID-19 diagnoses, based on linked official data on PCR tests. This dataset helped to create a clear picture of how ethnicity and deprivation affect COVID-19 risk. As the UK Biobank is not a representative sample of the UK population,[12] absolute risks should be interpreted carefully. However, the adjusted relative risks shown here provide an unambiguous signal of the disparities in COVID-19 incidence across the UK.

In conclusion, we found a strong association between ethnicity and the risk of COVID-19 infection in the UK, independent of other sociodemographic factors and comorbidity. In addition, socioeconomic deprivation and common chronic conditions (hypertension, obesity, diabetes, chronic respiratory disease, heart failure, and renal failure) were independently related to an increased risk of COVID-19 disease. Public health strategies to control the current pandemic should take these factors into account to best mitigate the spread of disease and mortality related to COVID-19 infection.

## Data Availability

Data can be accessed by application to UK Biobank (www.ukbiobank.ac.uk/register-apply). UK Biobank was established as a resource to allow scientists to carry out research into a wide range of diseases. The resource is available for researchers to use without preferential or exclusive access and all researchers, academic and commercial, are subject to the same application criteria, approval procedures and follow-up process.

## Transparency declaration

The lead author affirm that the manuscript is an honest, accurate, and transparent account of the study being reported; that no important aspects of the study have been omitted; and that any discrepancies from the study as planned have been explained.

## Ethical approval

The National Health Service’s National Research Ethics Service approved the collection and use of UK Biobank data.

## Acknowledgements

The authors acknowledge English language editing by Dr Jennifer A de Beyer of the Centre for Statistics in Medicine, University of Oxford.

## Contributorship statement

All authors contributed to the design of the study and interpretation of the results, and reviewed the manuscript. AP-U had access to the data, performed the statistical analysis, and acted as guarantor. AP-U, RP and DP-A wrote the first draft of the manuscript. DP-A is the senior author. The corresponding author attests that all listed authors meet authorship criteria and that no others meeting the criteria have been omitted.

## PPI statement

Patients and participants were and are involved in the UK Biobank study itself. No patients were involved in setting the research question or the outcome measures. Patients were not invited to comment on the study design and were not consulted to develop patient-relevant outcomes or interpret the results. Patients were not invited to contribute to the writing or editing of this document for readability or accuracy.

